# Revealing the Unseen Burden: A Comprehensive Analysis of Social and Economic Impact of Atopic Dermatitis in Argentina

**DOI:** 10.64898/2026.07.23.26358757

**Authors:** Natalia Espinola, Agustin Casarini, Natalí Ini, Asociación ADAR, Paula Luna, Constanza Silvestrini Viola, Federico Augustoski

## Abstract

Atopic dermatitis (AD) is a chronic inflammatory skin condition with a multidimensional burden. This study aimed to estimate the socioeconomic burden of the pediatric and adult population with AD in Argentina and explore the health system’s response to their needs. We employed a mixed-methods approach, combining a cross-sectional cost-of-illness analysis with a qualitative component to comprehensively assess the social and economic impact of atopic dermatitis in Argentina. We collected data from a survey, focus group and in-depth interviews conducted in Argentina between May 2023 and September 2023. We recruited adult patients and caregivers of children with AD below 18 years of age. We used multivariable ordinal regression to assess associations between sociodemographic and clinical characteristics and burden associated with AD. 1471 subjects participated in the pediatric analysis, and 1412 in the adult analysis. The average annual per capita cost was USD 2639 for adults with AD and USD 8682 for children with AD. Costs increased significantly with disease severity in both groups and were strongly associated with patients’ quality of life. Patients and caregivers highlighted the poor response of the health system in terms of adequate and timely access, as well as comprehensive care of the disease. Patients and caregivers suffer from a significant economic burden and quality of life impact associated with AD, which requires urgent attention and comprehensive support from the healthcare system to improve their lives.

## Introduction

Atopic dermatitis (AD) is a chronic inflammatory skin condition characterized by intense itching that affects 10-15% of the population, with a higher prevalence in children (20%) than in adults (1-3%) (1–3). In Argentina, the prevalence of AD in individuals aged 6-7 years and 13-14 years is 6.4% and 7.2%, respectively, while the AD prevalence in adults is 5.3% (4).

Based on Global Burden of Disease data, AD has the highest burden among all skin diseases, making it a significant public health problem worldwide. (5) AD patients lead to skin pain, sleep disruption, mental health issues, impaired quality of life, and a multidimensional burden encompassing physical, emotional, psychological, social, and economic challenges, particularly among patients with more severe disease, representing a significant burden for both patients and their families (6–11). In addition, a significant economic burden is associated with AD in terms of direct medical costs for the health system (such as doctor visits, hospitalization, and medication) and societal costs (such as out-of-pocket expenses, loss of work productivity, caregivers’ time, and reduced quality of life) (12). Several international studies have estimated the economic burden associated with AD, mainly in European countries, the United States, and Asian countries. However, this information is lacking in Latin American countries. According to the international literature, the economic burden associated with AD comprises approximately 30% direct medical costs and 70% other societal costs, depending on the level of disease severity (13,14). The main component of societal costs is the loss of work productivity of the patient and/or caregiver, followed by out-of-pocket expenses. More than 75% of the out-of-pocket costs resulted from household items, emollients, and medications (13). In the United States, the national cost of AD ranged from 364 million to 3.8 billion dollars in 2008, reflecting the challenges in thoroughly estimating the socioeconomic burden due to variations in cost components, data sources, and methodological approaches used in different studies (15). A study showed that adults with AD in the United States have an unadjusted annual healthcare incremental cost of $4979 compared to adults without AD in 2018, driven by outpatient services and pharmacy (16). In Europe, the average annual cost per adult with AD was between 2242 and 6924 euros in 2017 (14).

Furthermore, AD has a substantial negative impact on health-related quality of life (HRQoL) and more significant psychological distress than the general population and individuals with other conditions (6,8,14,17,18). In addition, several studies have reported a lower quality of life among families with children with AD compared to those with children without AD (6). A study conducted in the Latin America and Caribbean region, specifically in Colombia, reported that the most affected component in the quality of life measure (EQ-5D) was the presence of pain/discomfort and anxiety/depression, intensifying the symptomatic and emotional impact of the disease (19). As for Argentina, there is limited evidence regarding the effects of AD on costs for patients and caregivers, as well as on psychological comorbidities. The available evidence consists of unpublished studies conducted by an AD patient association in Argentina (20).

The Argentine health system is characterized by decentralization in the public sector and fragmentation in social insurance, both in funding sources and service provision. The public subsector is financed at national, provincial, and municipal levels, with provinces having autonomy over health strategies and resource management. It provides healthcare services mainly to the uninsured through public hospitals and primary care centers. Social security covers salaried workers, retirees, and public employees, though many are financially inefficient due to small risk pools. The private sector relies on out-of-pocket payments and prepaid insurance, dominated by a few firms. The Argentine health system is characterized by high fragmentation due to the lack of effective coordination and cooperation mechanisms between the three subsectors regarding management, financing, and health risks (21). According to the 2022 Census, 60.9% of the population has social security or private coverage, while 39.1% lacks formal health coverage (22).

In this regard, the fragmentation and decentralization in the provision of healthcare services within Argentina’s healthcare system, along with the socioeconomic inequalities in the country, can result in unmet healthcare needs for individuals with AD and become a significant burden for some population groups. However, to date, the socioeconomic burden of AD remains inadequately understood within the Latin American region.

The objectives of this study were to estimate the socioeconomic burden associated with AD in Argentina and to explore patients’ and caregivers’ experiences with the health system response. We also investigated the associations between costs, sociodemographics characteristics, HRQoL, and the severity of AD through a multivariate econometric model.

These findings can help better understand the impact of AD on Argentinian patients, healthcare systems, and the overall society to improve its management.

## Methods

### Study design and sample selection

Our study had two main components and used mixed research methods: 1) a quantitative cross-sectional cost-of-illness survey to assess the economic and quality-of-life burden attributable to AD and 2) a qualitative component to explore how patients and caregivers understand their experiences with this chronic disease, and how the healthcare system addresses their needs.

We conducted online cross-sectional surveys for two target populations in Argentina: 1) adults (aged 18 years and above) diagnosed with AD and 2) pediatric population with AD through their respective caregivers. We employed a convenience sampling method, a nonprobabilistic and nonrandom sampling technique. The dissemination and data collection period lasted four months, from June 2023 to September 2023.

We used a qualitative methodological design based on focus groups and semi-structured interviews. We recorded the focus group and interviews, transcribed them verbatim and anonymised them to ensure the confidentiality of the participants. We uploaded the transcripts into the software Atlas.ti. We guided an initial list of codes on literature and discussion with the patient association, complemented by emerging codes. We analyzed the data following a framework analysis approach. Results were discussed with the patient association and the entire team.

The sample was purposive to seek representativeness in the severity spectrum of AD, subnational level, and healthcare sector within the health system in Argentina (social security, private sector, and public sector). Recruitment was performed with the support of the patient association Argentinian Atopic Dermatitis Civil Association (ADAR, by its Spanish acronym).

ADAR is a legally constituted non-profit civil association composed of patients and relatives of patients with AD. The association, founded in 2018, aims to contribute to improving the quality of life of people affected by atopic dermatitis, promoting education, and promoting research in Argentina.

### Survey development and adaptation

We developed an online self-administered survey based on established international questionnaires (23,24). The first questionnaire was developed for patients aged 18 years and above diagnosed with AD and covered the following topics: sociodemographic and clinical data, out-of-pocket expenses related to AD management, employment status, work productivity, and quality of life. The second questionnaire developed for pediatric patients (0 to <18 years) with AD and caregivers covered the following topics: sociodemographic data of the caregiver and child, caregiver’s employment status, caregiver’s work productivity, time spent on caregiving, quality of life, clinical data, loss of school days, and out-of-pocket expenses related to AD management. The questionnaires are in section 1 of the supporting information (SI).

The interview and focus group guides covered the following topics: barriers and facilitators in the diagnosis phase; the impact of the disease on daily life at work, school, and emotional levels; and obstacles, unmet needs, and possible improvements in the relationship with the healthcare system. The interview and focus group guides are in section 2 of the SI.

### Measures and variables

#### Sociodemographics and clinical characteristics

The respondents answered several sociodemographic questions, including province, age, gender, health coverage, educational level, and employment status. They also reported clinical data, including the severity of AD and whether they had flare-ups during the last month.

#### Out-of-pocket expenses

The out-of-pocket expenses included expenses in services and/or products for managing AD, such as moisturizers, out-of-pocket payments for medical visits, hospitalization and certain medications (such as corticosteroids), transportation expenses, and personal hygiene. Out-of-pocket expenses were reported as a monthly cost per item and converted into an annual average cost, assuming they had similar monthly expenses.

#### Cost of lost work productivity

We estimated the productivity loss from absenteeism and reduced productivity at work due to illness (presenteeism). Absenteeism hours were self-reported weekly and extrapolated annually, assuming a constant absenteeism pattern. We also estimated the maximum working hours to be 48 hours per week. The percentage of lost work hours was calculated by dividing the self-reported lost hours by the average hours worked by individuals with similar demographic information. These last data are presented in Section 3, Table 2 in SI. In addition, we collected information on the percentage of impairment while working due to health problems when the individual due to AD cannot work at 100% capacity. The total cost of lost productivity was valued using the human capital approach, including the average hourly wage for absenteeism hours and the hourly proportionate salary for reduced productivity at work (presenteeism). The data on hourly wage gender, age, and educational level was collected from the second quarter of the 2023 Permanent Household Survey (EPH) conducted by the National Institute of Statistics and Censuses (INDEC, in Spanish) (25). Section 3, Table 2 in SI, presents the hourly wage data.

**Table 1.**
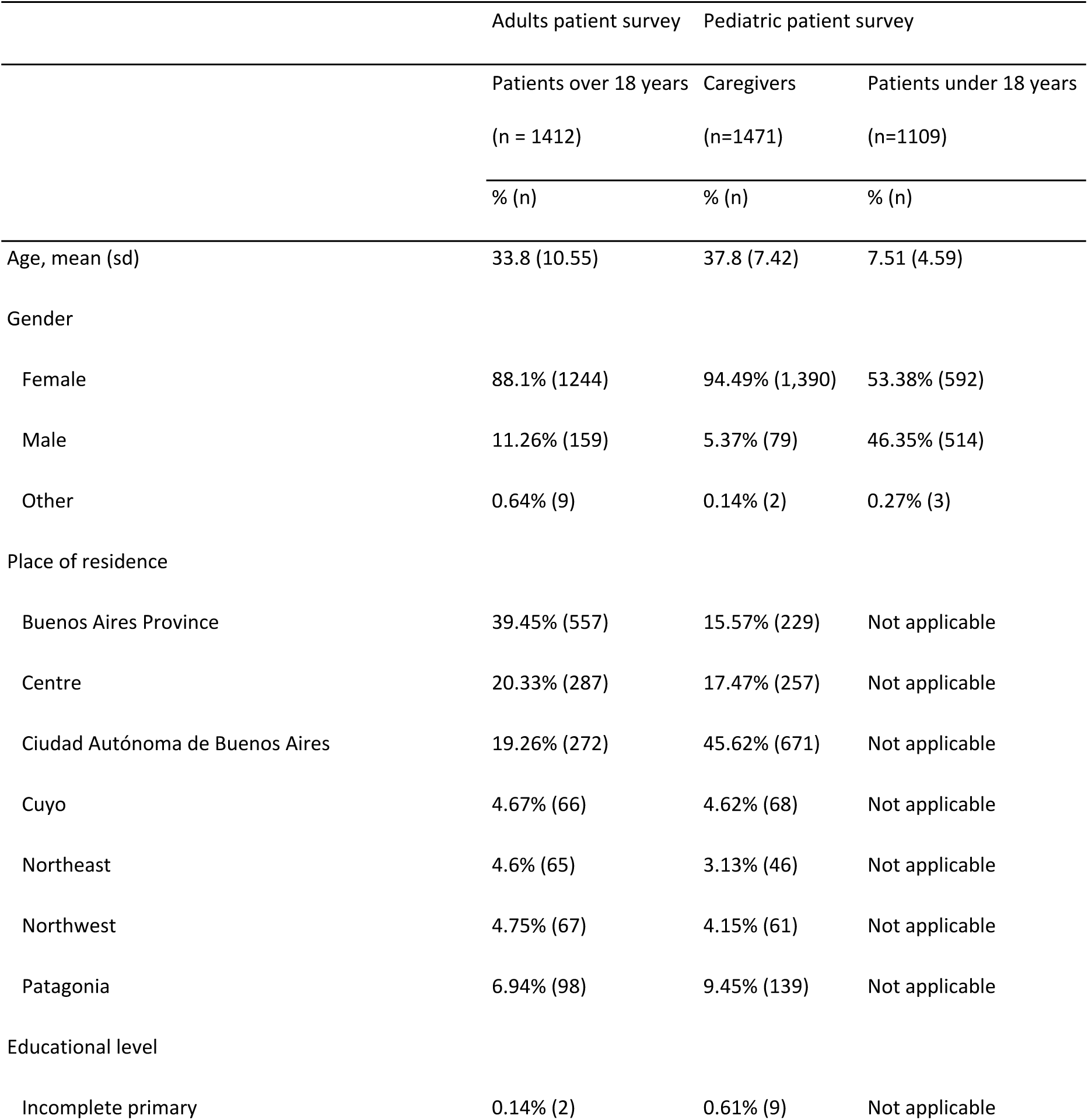
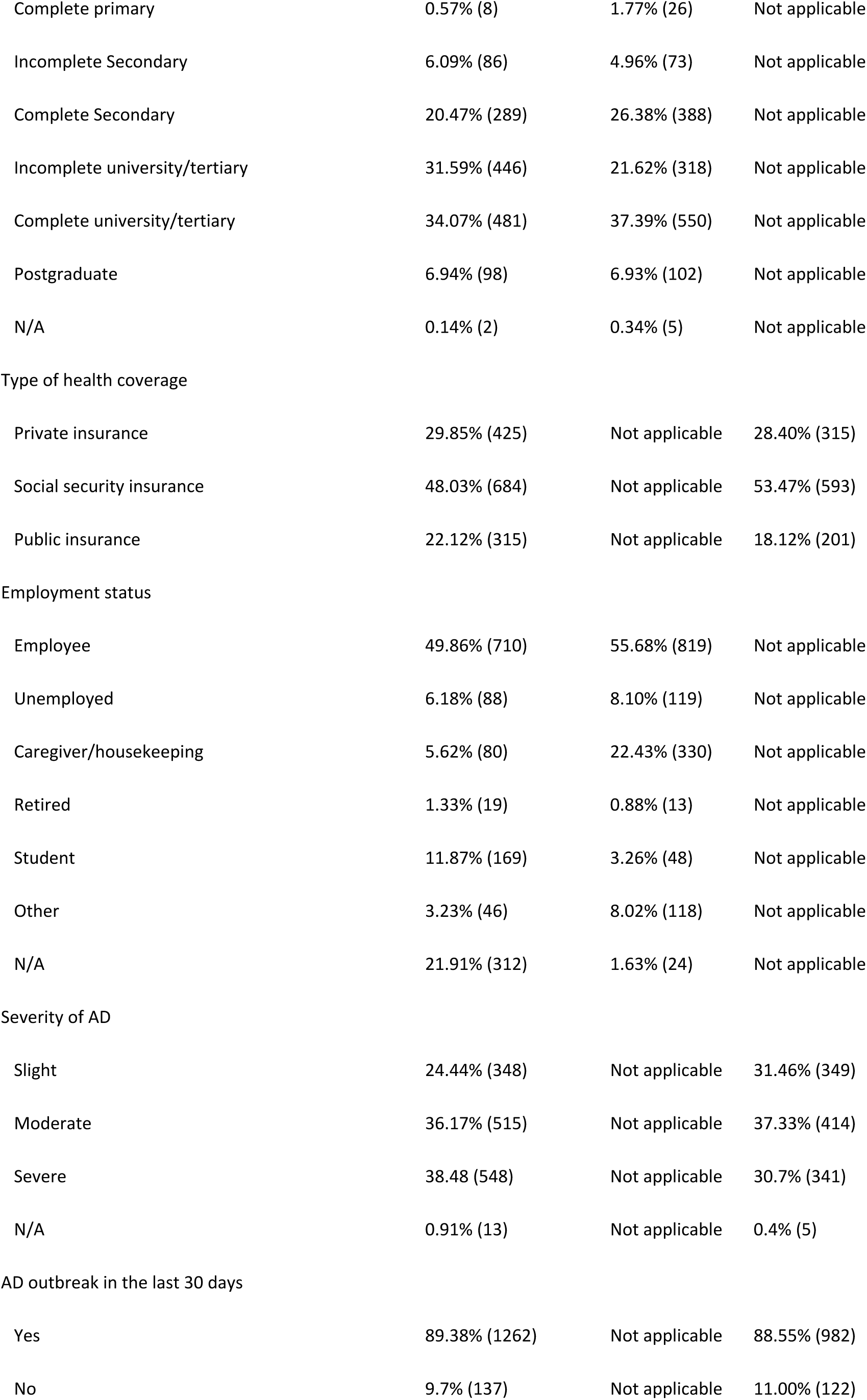
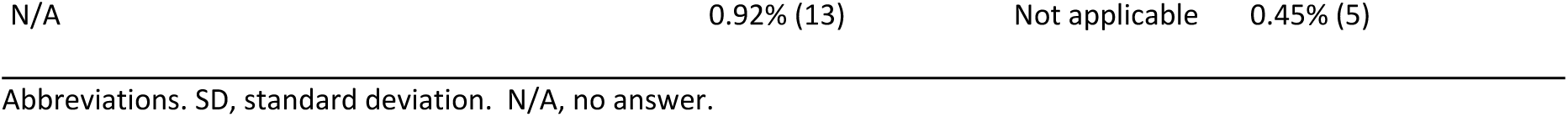
Descriptive statistics of sample.

**Table 2.** Average out-of-pocket expenditure and productivity cost per AD adult patient and severity, expressed in USD 2023.

| Cost Component | N | AD severity |  |  |  |  |  | Difference<br>between<br>mild and<br>severe<sup>1</sup> | Average<sup>2</sup> |
| --- | --- | --- | --- | --- | --- | --- | --- | --- | --- |
|  |  | Mild |  | Moderate |  | Severe |  |  |  |
|  |  | Mean | sd | Mean | sd | Mean | sd |  |  |
| Topical treatments<sup>3</sup> | 1,102 | \$328 | \$540 | \$410 | \$410 | \$535 | \$699 | \$207*** | \$378 |
| Drugs | 1,106 | \$99 | \$380 | \$132 | \$195 | \$304 | \$822 | \$205*** | \$129 |
| Private Doctor/Visit to<br>Emergency Department | 1,103 | \$93 | \$193 | \$120 | \$225 | \$284 | \$1229 | \$191** | \$119 |
| Hospital admissions | 1,105 | \$3 | \$32 | \$2 | \$24 | \$8 | \$60 | \$5* | \$3 |
| Travel expenses | 1,103 | \$21 | \$98 | \$27 | \$97 | \$69 | \$367 | \$48** | \$28 |
| Personal hygiene<sup>4</sup> | 1,106 | \$154 | \$226 | \$190 | \$260 | \$287 | \$555 | \$133*** | \$179 |
| Other | 1,110 | \$75 | \$255 | \$91 | \$287 | \$256 | \$1061 | \$181** | \$96 |
| Total out-of-pocket<br>expenditure | 1,116 | \$770 | \$1014 | \$967 | \$928 | \$1713 | \$3605 | \$942** | \$925 |
| People working (%) | 1112 | 68% | 47% | 67% | 47% | 59% | 49% | -9%** | 67% |
| Work absenteeism, annual<br>hours | 691 | 51 | 247 | 120 | 409 | 301 | 656 | 252*** | 99 |
| Working hours lost (%)<sup>5</sup> | 691 | 3% | 13% | 5% | 18% | 14% | 29% | 11%*** | 5% |
| Adjusted Presenteeism (%)<sup>6</sup> | 691 | 24% | 24% | 40% | 28% | 56% | 29% | 32%*** | 33% |
| Work impairment (%) | 691 | 25% | 26% | 42% | 30% | 59% | 30% | 34%*** | 35% |
| Cost of lost productivity due<br>to absenteeism | 691 | \$201 | \$792 | \$572 | \$2027 | \$1333 | \$3124 | \$1132*** | \$442 |
| Cost of lost productivity due<br>to presenteeism | 691 | \$1378 | \$427 | \$2666 | \$992 | \$4153 | \$2130 | \$2775*** | \$2127 |
| Total cost of lost<br>productivity | 691 | \$1579 | \$782 | \$3238 | \$1875 | \$5486 | \$2642 | \$3907*** | \$2570 |

| The average total social cost | \$1842 | \$3126 | \$4938 | \$2639 |
| --- | --- | --- | --- | --- |

<sup>1</sup> p-value: \* <0.10, \*\* <0.05, \*\*\*<0.01. <sup>2</sup> The average was obtained using the prevalence by the severity scale POEM from Barbarot (2018) in Spain. The prevalence weights are for mild 50.6%, for moderate 41.6%, for severe 7.6%. <sup>3</sup> For example, moisturizers, topical steroids, and topical calcineurin inhibitors. <sup>4</sup>For example: soaps, syndets, shampoo for atopic skin, soap-free soap, other specific cleaning products for atopic skin, etc. <sup>5</sup> Estimated by dividing the self-reported hours lost by the average number of hours worked for people of the same age, gender, and educational level obtained from the EPH conducted by the INDEC. <sup>6</sup> The presenteeism percentage was calibrated based on a literature review explained in the data validation section. <sup>7</sup> The social cost is calculated by adding the average out-of-pocket expenses and the cost of lost productivity adjusted by people working.

#### Informal caregiving cost

Time spent on care tasks was self-reported weekly and extrapolated annually under the assumption of consistent caregiving routines. The maximum time allocated for care was estimated to be 16 hours per day, accounting for 8 hours of sleep per day. Informal caregiving costs were determined using the opportunity cost method (23), in which the number of hours spent caregiving was multiplied by the average hourly wage for working-age individuals of similar gender, age, and education level. For retired caregivers, a proxy good method was applied (23), using hourly wage rates based on the market (labor) prices of a ‘close market substitute’. We then assumed the average hourly wages of those who worked in health-related social assistance. This data is presented in Section 3, Table 3 in SI.

**Table 3.**
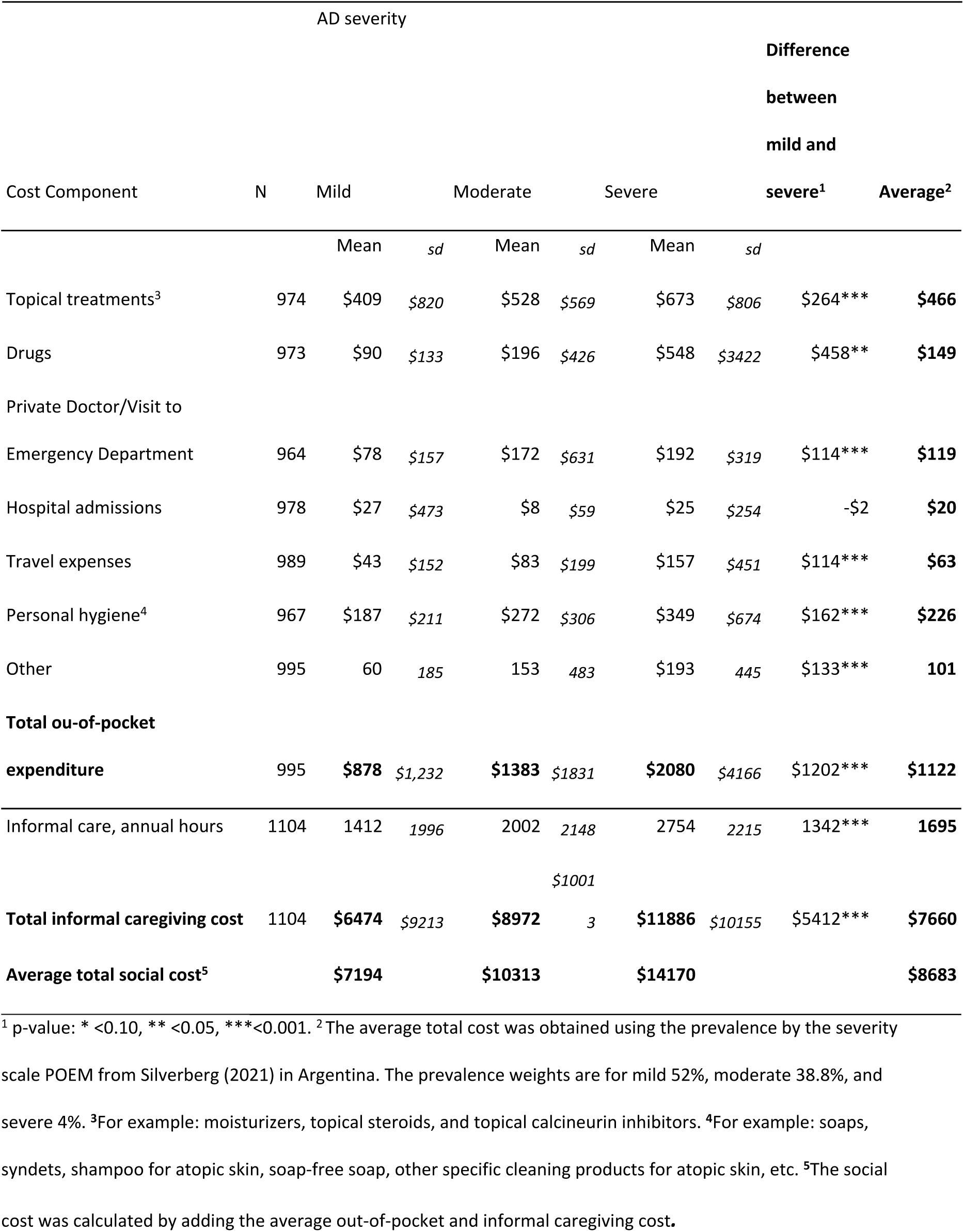
Average annual out-of-pocket expenditure and care cost per pediatric patient in USD 2023.

#### Estimation of Quality of life

The quality of life of adults with AD was assessed using the EuroQoL (EQ-5D-5L), which evaluates five health dimensions and includes a visual analog scale (EQ VAS) (26,27). For children, we used the child-friendly proxy version (EQ-5D-Y proxy version 1), where caregivers rated the child’s health-related quality of life (28). Informal caregiving-related quality of life was measured using the CarerQol, which includes a multidimensional assessment (CarerQol-7D) and a well-being scale (CarerQol-VAS) (29–32). Given the absence of national preference weights for Argentina, societal preferences from Uruguay (EQ-5D-5L), Spain (EQ-5D-Y), and the United States (CarerQol-7D) were used (26,33). See section 4 in the supporting information (SI) for more details.

#### Data analysis

The questionnaire’s database was processed and statistically analyzed using Stata® v17. The results are presented as the mean, standard deviation (SD), or percentage, depending on the variable’s distribution. The relationship between variables was evaluated through multivariate linear regression. In all cases, differences with a significance level of p≤0.05 were considered statistically significant.

In the qualitative analysis, inductive inquiry was used, consistent with the grounded theory approach. Data analysis was performed following the principles of grounded theory (34). The collected data were coded following a coding manual elaborated from the axes of the interview guide and emerging themes (35).

The average social cost per adult with AD was estimated by combining out-of-pocket expenses and productivity losses, adjusted for employment rates. For children, it included out-of-pocket expenses and caregiving costs. Severity-weighted means were applied to minimize bias using POEM-based severity distributions (Spain for adults, Argentina for children) (36,37). Moderate to severe and severe categories were merged for more explicit analysis and comparability with other studies.

Since the survey was conducted over four months amid high price variation in Argentina, out-of-pocket expenses were adjusted to September 2023 using the Consumer Price Index (38). Costs were initially reported in Argentine pesos (ARS) and converted to US dollars (USD) at the exchange rate of 1 USD = 365.60 ARS as of September 30, 2023 (39).

#### Calibration

An internal and external validation process was conducted to ensure accuracy. The research team reviewed the methodology, and a literature review was performed on key parameters like productivity and caregiving hours. Experts identified potential bias in presenteeism data for mild AD cases, possibly overestimating lost productivity costs. A calibration process adjusted presenteeism-based costs using literature findings by severity to address this. Details are provided in section 5 of the SI.

#### Ethics statement

This study was approved by Comité de Ética Central del Ministerio de Salud de la Provincia de Buenos Aires. Adults and caregivers received written information about the purpose of the study, and participation was voluntary before giving their consent to participate.

## Results

### Respondents characteristics

Out of 1670 surveys received from adults with AD, 258 were excluded due to incomplete sociodemographic data, resulting in a sample of 1412 adults with AD (Table 1).^1^ We compared sociodemographic characteristics between respondents who completed the survey in full and those who completed it only partially and found no evidence of demographic bias between the two groups (S3 Table 1). Their average age was 34 years (SD = 10.55), with 88% female and 40% residing in Buenos Aires Province. Over two-thirds had a complete or incomplete university education (66%), while 20% had a complete secondary-level education. Most participants had some form of health coverage, predominantly social security insurance (48%). Most adults had severe AD (38%), followed by moderate AD (36%). Furthermore, the vast majority (89%) reported experiencing a flare episode in the last 30 days.

Out of 1782 surveys received from caregivers who completed the information for the pediatric population, 311 were excluded due to incomplete sociodemographic details, resulting in a sample of 1471 caregivers and 1109 children (Table 1).^1^ The caregivers had an average age of 38 years (SD = 7.42), with 95% female and 46% residing in Buenos Aires Province. Educational level distribution is similar to that of adult patients. Over half were employed, while 22% engaged in family and household care tasks. The children under their care had an average age of 8 years (SD=4.59), with 53% being girls and 82% having health insurance. Most children had moderate AD (37%), followed by mild (32%) and severe (31%). Most patients experienced a flare in the last 30 days, just as adults did.

### Cost estimation for atopic dermatitis adults

On average, adults with AD spent USD 925 out-of-pocket in the year. The main cost category was Topical treatment (moisturizers, topical steroids, and topical calcineurin inhibitors), which represented 41% of the average expenses, followed by personal hygiene (19%). Working adults reported losing 99 hours in a year (1.9 hours/week) due to AD, representing 4.56% of the working hours per week on average in Argentina. In addition, working adults reported a 33% loss of their productivity at work due to AD. Therefore, the total cost of lost work productivity for working adults with AD was estimated at USD 2570. Therefore, the average social cost per adult with AD in one year was estimated at USD 2639. This cost increases with disease severity. The cost difference between adults with severe and mild AD is approximately three times (p-value < 0.01). See Table 2. More detailed information is available in the Section 6 in SI.

### Cost estimation for the pediatric population with AD

On average, children with AD required out-of-pocket expenditures of USD 1222 per year. Similar to adults with AD, the main expense category was topical treatment (41%). Out-of-pocket expenses increase in children with public insurance, while this phenomenon has not been observed in adults with AD (See Section 7 in SI). In addition, caregivers reported dedicating 1695 hours a year (33 hours/week) on average to the healthcare of their children with AD, representing an average cost of USD 7660. Therefore, the average total cost per pediatric patient with AD was estimated at USD 8683.

On the other hand, caregivers also reported work productivity losses; the results of these estimates can be seen in Table 4 of Section 8 of the SI. In addition, caregivers reported that 21% of children with AD had missed school in the last month, averaging eight days per year, representing a 4% loss of the academic year. The cost and number of missed school days attributable to AD increased with disease severity. See Table 3. More information is available in Section 8 of the SI.

**Table 4.** Systematized qualitatives findings on the experiences of patients with atopic dermatitis and their caregivers.

| Category | Details |
| --- | --- |
| Diagnosis and disease course | Symptoms at birth or infancy, intermittent condition, daily care, moisturizers, periodic peaks |
| Treatment | Topical steroids, corticosteroid cream, IV corticosteroids for severe cases, biological therapies for significant improvement |
| Access to healthcare and medication | Difficulty securing appointments, private insurance access, out-of-pocket for private healthcare, dissatisfaction with emergency healthcare |
| Doctor-patient relationship | Lack of information about AD, prescribed topical treatments, felt judged, symptoms minimized, conflicting opinions between dermatologists and allergists, appreciated referrals to specialists |
| Costs | Economic costs linked to moisturizers, topical treatments, out-of-pocket payments, creams not covered by insurance, biological medications only accessible in private insurance |
| Searching for information | Internet searches for treatments and causes, scarcity of information from health professionals, valued peer recommendations |

### Psychosocial burden of atopic dermatitis

Figure 1 shows the health-related quality of life (HRQoL) measured using the EuroQol tools for adult AD patients and children. In the first case, physical and mental problems were predominant (79% and 86%, respectively). In the last case, most reported no mobility or personal care problems (94% and 68%, respectively); however, half of the participants reported some difficulties in most categories. In summary, for adults with AD, the mean (SD) EQ-5D-5L index was 0.87 (0.11), and the VAS score was 62.29 (0.67), while for children with AD, EQ-5D-Y was 0.69 (0.24) and the VAS score was 67.34 (22.74). The multivariate analysis showed that quality of life in adults with AD declines with increased severity, higher productivity loss, male gender, and public vs. private health coverage (see Section 9 in the SI). In children, lower quality of life was associated with social security or no health coverage vs. private insurance, greater disease severity, and higher caregiving-related social costs (see Section 10 in the SI).

**Figure 1.**
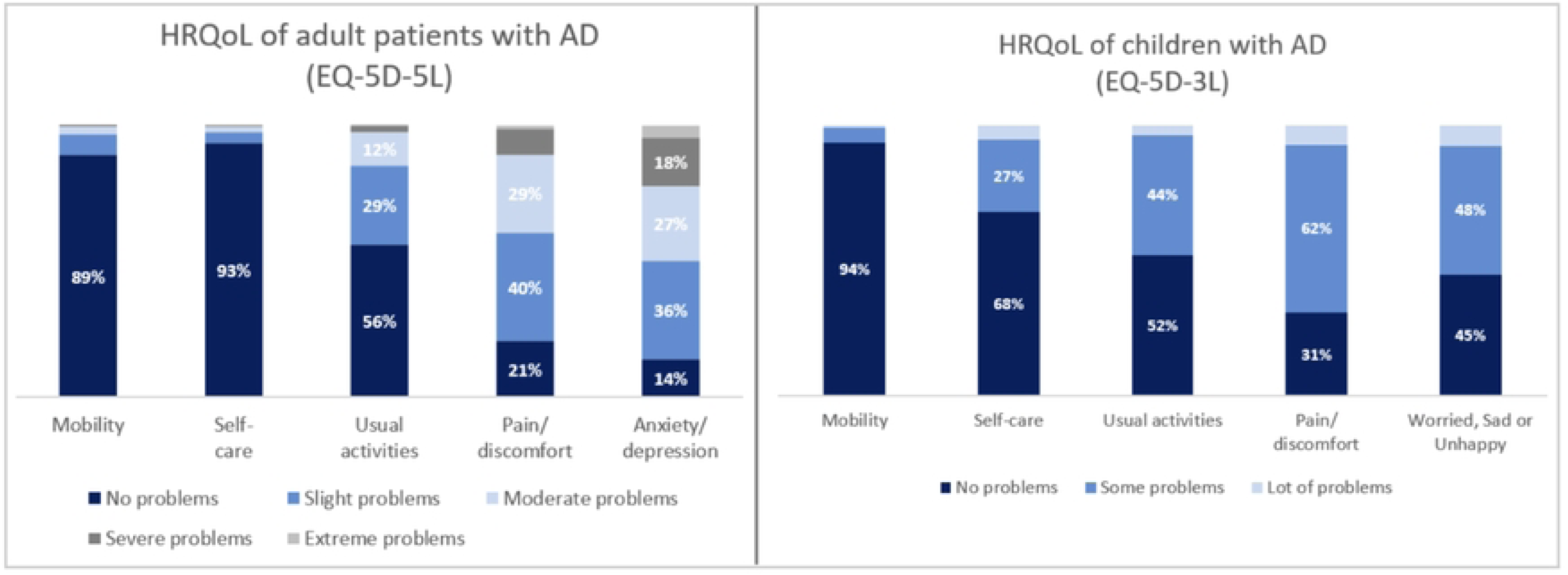
HRQoL of adults and children with atopic dermatitis.

Figure 2 shows the caregivers’ quality of life measured using the CarerQol-7D descriptive system. Approximately 60% of the respondents claimed to have financial, physical, and mental problems. 52% reported feeling very satisfied with their caregiving tasks, while 43% reported lacking support in their caregiving duties. In brief, the mean (SD) CarerQol-Tariff was 45.89 (19.64), while the CarerQol-VAS score was 60.45 (27.64).

**Figure 2.**
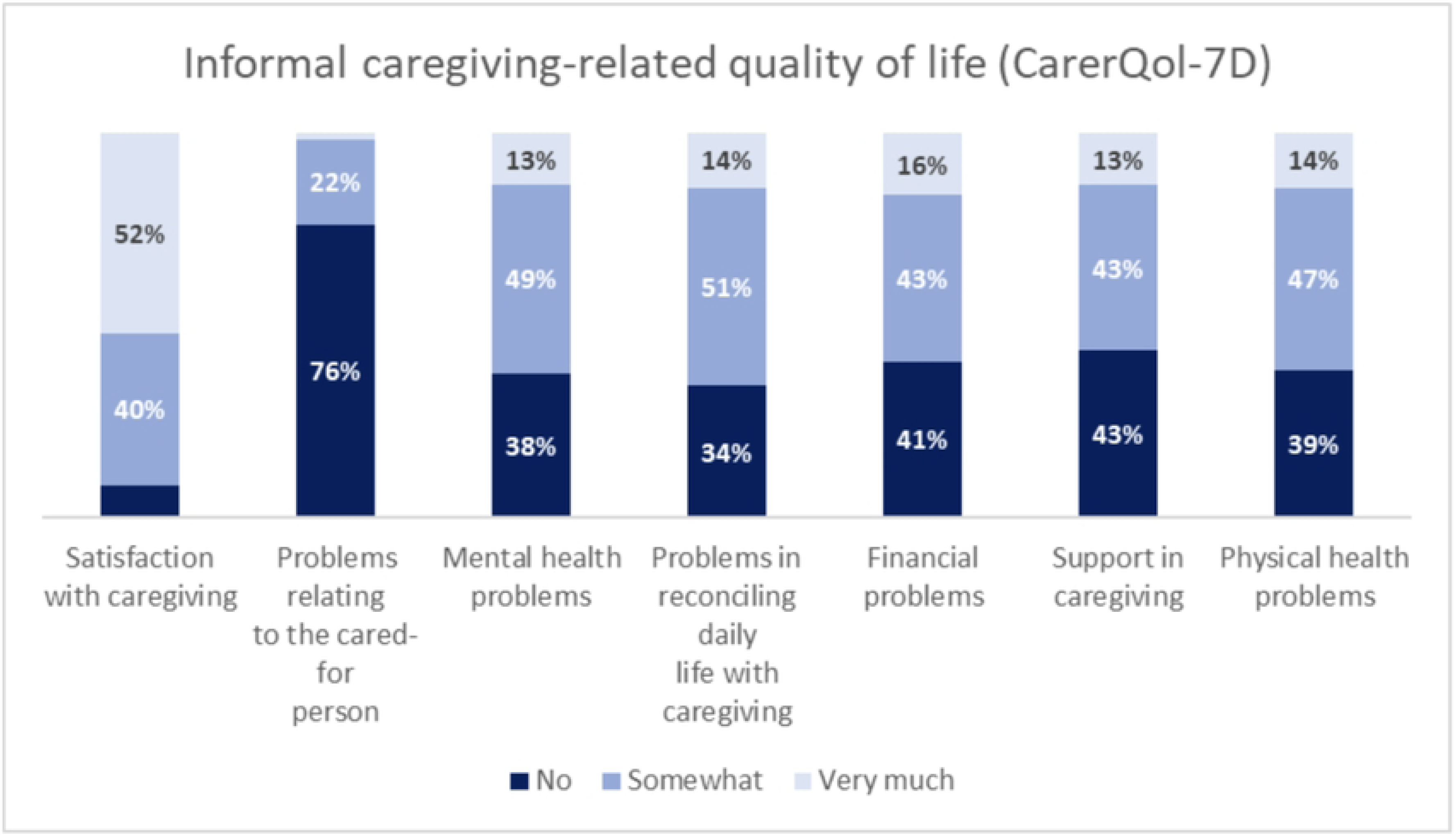
Quality of life related to informal caregiving

### Qualitative findings. Analysis of the health system’s response to unmet needs

The main findings from qualitative research were systematized in Table 4. Participants reported a delayed diagnosis of AD, with symptoms typically starting from birth or early childhood. Some caregivers discovered their AD after their children were diagnosed. The disease was characterized by intermittent flare-ups, requiring continuous care and the use of moisturizers. Topical steroids were commonly used for mild cases, while severe cases often required IV corticosteroids. A few participants had access to biological therapies, which they found effective in improving their condition. However, access to healthcare was challenging, especially for those without private insurance, who struggled with long wait times and high out-of-pocket costs for treatments and specialist consultations.

The doctor-patient relationship was a concern, as many participants felt they were not provided with adequate information about AD and often felt judged for not managing the condition properly. There was also a lack of coordination between dermatologists and allergists, leading to conflicting treatment approaches. Financial burdens were significant, with moisturizers and topical treatments often not covered by insurance and biological medications only accessible through private insurance. Participants expressed frustration over the lack of information from healthcare professionals and frequently turned to the Internet or peer support for advice and information on treatments and causes of AD.

Regarding the impact of the disease on daily activities, the key points mentioned by participants include:

- Need for comprehensive and multidisciplinary treatment, including psychological support.
- School attendance hindered by the emotional and physical effects of the disease.
- Significant impact on their work lives. Required to cease employment or adjust. Dismissals due to absenteeism, need to obtain a disability certificate, hostility in the workplace due to absenteeism, and physical appearance.
- Feelings of anxiety, anguish, depression, and insomnia, which were primarily attributed to discomfort resulting from the disease and the visible symptoms thereof.
- Difficulties in social interactions owing to physical appearance.
- Caregivers reported a state of alertness and fatigue.

For more details on the qualitative findings, see Section 11 in SI.

## Discussion

Our study is the first in Argentina and the Latin American region to estimate the economic and social burden of atopic dermatitis (AD) through primary data collection, employing both quantitative and qualitative research methods and incorporating adult and pediatric patients and caregivers. Our findings showed that the average societal annual cost associated with AD was USD 2639 per adult (USD 1842 for mild AD vs. USD 4938 for severe AD, p<0.001) and USD 8683 per pediatric patient (USD 7194 for mild AD vs. USD 14170 for severe AD, p<0.001).

Adults and pediatric adults with AD have spent approximately USD 1000 annually on average out-of-pocket, representing 7% and 9% of the average income, respectively. Both adult patients and caregivers allocated most of their out-of-pocket expenses to topical treatment (around 40% of the total expenditure). Similar findings have been reported in other countries (13,40). These products are categorized as cosmetic items in Argentina and are not covered or discounted. This point is reinforced by our findings, which indicate that the absolute value of out-of-pocket spending does not vary significantly across different types of health coverage. However, a key concern is the financial burden this spending represents relative to income. Caregivers and patients relying on public insurance generally have lower incomes, making these out-of-pocket expenses a significant financial constraint, potentially exacerbating healthcare inequities.

The findings also show that AD imposes a significant burden on labor productivity. Adults with AD lost approximately 5% of their annual working hours due to absenteeism and reported a 33% reduction in productivity due to presenteeism. These results align with previous studies (14,41–45). The qualitative data corroborate these findings, with job stability emerging as a significant concern. Many participants reported difficulties in maintaining employment, securing jobs, and experiencing dismissals due to absenteeism. In addition, caregivers dedicated approximately 5 hours per day to care for affected individuals, representing an annual economic burden of USD 7659 per informal caregiver. This estimate is higher than findings from high-income countries such as Australia, where parents reportedly spent between 2 and 3 hours daily caring for a child with AD (6,46). This difference may be attributed to the limited availability of formal caregiving services in Latin American countries compared to high-income settings, where structured support systems help alleviate caregiving responsibilities for families. The lack of formal caregiving services not only increases the burden on informal caregivers but also exacerbates the financial strain on families. Many caregivers and adults with AD must reduce their working hours or leave their jobs entirely, leading to a significant loss of household income.

The average estimated EQ-5D-5L index for adults with AD in our study was 0.87, aligning with previous research and comparable to indices reported for conditions such as diabetes and arthritis (46,47). Focus groups and interviews further highlighted high levels of anxiety, distress, depression, and insomnia among both patients and caregivers, often due to discomfort and the visible symptoms of AD. The constant vigilance required by caregivers negatively affected their sleep and overall well-being, contributing to chronic stress and fatigue. These findings underscore the broader psychosocial burden associated with AD and highlight the need for comprehensive support systems that address not only medical but also emotional and mental health needs.

Given that AD is not classified as a chronic disease in Argentina—except in the province of Misiones—this classification gap can further limit access to insurance coverage, work-related benefits, and reimbursement for essential treatments. The absence of comprehensive coverage further exacerbates the financial burden on families, reinforcing health inequities and the need for policy reforms to ensure adequate support for individuals living with AD. In September 2023, Misiones enacted the Law of Atopic Dermatitis Care (Provincial Law No. 1700), establishing a comprehensive approach to the disease. This law mandates early detection, diagnosis, and treatment while ensuring multidisciplinary medical care and access to pharmacological and therapeutic interventions. It also promotes telemedicine for remote patient assistance and follow-up, as well as research and education on AD. The law provides coverage for diagnostic tests, psychological and rehabilitative therapies, phototherapy, and essential medications, including biologics and emollients. This initiative could serve as a precedent for national-level policies to improve AD management and access to care in Argentina.

Our study has several strengths, including developing a questionnaire validated by experts. We also employed a qualitative methodology to capture complexities in the relationships between AD patients, caregivers, and the healthcare system. We engaged a skilled qualitative researcher to moderate focus groups, potentially fostering open expression from informants. However, this study has some limitations. While the achieved sample size in the quantitative research exceeded the initial goals, the sample was not randomly selected, potentially affecting the representativeness of the results. Some questions required complex recall, introducing the potential recall bias inherent in time use and cost research. Although standardized questionnaires would have been preferred for measuring disease burden, survey format adherence might not have been absolute because it prioritizes a reduced question count to improve response rates. Adapted questions for presenteeism loss initially yielded higher values, which were corrected through a literature review. The adjusted productivity loss relied on salary estimates from national household surveys, potentially not fully representing the sample.

Additionally, using general quality-of-life questionnaires rather than specific AD questionnaires may not capture particular aspects of the disease, although it allows comparisons with other diseases. Another limitation of this study is the sampling bias inherent in recruiting participants through patient associations, which resulted in an overrepresentation of severe cases and female participants. This bias likely reflects that individuals with more severe disease and women are more proactive in seeking support through such associations, as evidenced by studies on gender differences in healthcare access. To mitigate this bias, cost estimates were adjusted using weighted averages based on severity distributions reported in the literature.

It is important to note that the socioeconomic burden of AD may be underestimated, as this condition is associated with various comorbidities such as allergic rhinitis, pollen allergy, dust allergy, bronchial asthma, and other dermatological diseases (46). Another point to highlight is that many AD adults are also caregivers for children with AD, which could significantly intensify the economic burden. Finally, it is vital to highlight the importance of multidisciplinary work with the community affected by AD and an integrated approach by healthcare professionals.

## Conclusion

The socioeconomic burden of AD in Argentina implies not only a significant impact on health-related quality of life but also a high cost for the health system, patients, and caregivers. In this regard, public policies are needed for patients with AD and caregivers.

## Data Availability

Data cannot be shared publicly due to the sensitive nature of the health information collected and the terms of the informed consent obtained from participants, which did not include consent for public data sharing. This restriction was set by the Comité de Ética Central del Ministerio de Salud de la Provincia de Buenos Aires (approval number: ACTA-2022-36100592-GDEBA-CECMSALGP). Interested researchers may request access to the minimal dataset by contacting the corresponding author, Natalia Espinola, subject to approval by the ethics committee and execution of a data use agreement.

## Acknowledgments

We thank Pfizer for the funding provided to develop this project.

## Abbreviations

AD: Atopic dermatitis
ADAR: Argentine Civil Association of Atopic Dermatitis
ARS: Argentine pesos
CarerQol: Care-related Quality of Life
CarerQol-7D: Care-related Quality of Life, 7 Dimensions
CarerQol-VAS: Care-related Quality of Life Visual Analogue Scale
EQ-5D-5L: EuroQol 5-Dimension 5-Level
EQ-5D-Y: EuroQol 5-Dimension Youth
EQ VAS: EuroQol Visual Analogue Scale
EPH: Permanent Household Survey
HRQoL: Health-related quality of life
IECS: Institute for Clinical Effectiveness and Health Policy
INDEC: National Institute of Statistics and Censuses
POEM: Patient-Oriented Eczema Measure
SD: Standard deviation
SI: Supporting information
USD: United States dollars

